# Impact of Seasonal Variability on Cerebrovascular and Cardiovascular Health: Evidence from a District Hospital in Ghana

**DOI:** 10.64898/2026.07.28.26359183

**Authors:** Akua A. Karikari, Stephen A. Karikari, Lord B. Amponsah, Enoch Agbeleseshie, Victor Caesar, Akosua B. Karikari

## Abstract

The burden of cardiovascular and cerebrovascular diseases is rising rapidly across sub-Saharan Africa. Increasing scientific reports indicate that meteorological seasonality may be associated with vascular hospitalization, yet evidence from tropical West Africa remains scarce. Understanding how climate and seasonal factors influence vascular health and hospital admissions is essential for informing targeted, context-specific health system responses.

We conducted a hospital-based ecological time-series analysis using 45 months of electronic health records (February 2022 to August 2025) from Ga North District Hospital, Accra, Ghana (N = 8,308 admissions). Vascular burden was classified from primary discharge diagnosis into three mutually exclusive categories: cardiovascular, cerebrovascular, and non-vascular. Monthly climate data obtained from the Ghana Meteorological Agency (GMet) were linked by admission month and used to characterize the three meteorological seasons. Logistic regression examined associations between standardized climate variables and vascular admission. Multivariable models incorporated non-redundant climate variables and were adjusted for age and sex.

There were 427 cardiovascular admissions (5.1%) and 672 cerebrovascular admissions (8.1%) over the study period. Cardiovascular cases nearly tripled as a proportion of all admissions between 2022 (2.4%) and 2024 (7.1%). GMet data confirmed three climatically distinct seasons namely, Dry and Warm, Wet and Cool, Wet and Warm. Both diseases were disproportionately concentrated in the Wet and Cool season (cardiovascular 46.1%; cerebrovascular 42.1%; p < 0.001). Simultaneous adjustment for all non-redundant climate variables revealed contrasting effects of rainfall characteristics on cardiovascular admissions. While increased total monthly rainfall was associated with a lower likelihood of admission (adjusted OR 0.72, 95% CI 0.61–0.85, p < 0.001), a higher frequency of rainy-days was independently associated with an increased probability of admission (adjusted OR 1.73, 95% CI 1.38–2.18, p < 0.001). No climate variable independently predicted cerebrovascular admission. Older age independently predicted both cardiovascular and cerebrovascular admissions, whereas male sex specifically predicted cerebrovascular admissions (AOR = 1.30, 95% CI 1.10–1.52, p = 0.002).

The Wet and Cool season was associated with a higher concentration of vascular admissions at Ga North District Hospital, with rainfall frequency independently predicting cardiovascular admissions. Cerebrovascular admissions, in contrast, were more strongly associated with patient-level characteristics than with the climatic variables examined. These findings support seasonally informed resource planning at district facilities in tropical West Africa and underscore the importance of climate-resilient vascular health systems.

## Introduction

Cardiovascular and cerebrovascular diseases are the leading causes of death and disability globally, accounting for an estimated 18.6 million deaths in 2019 alone [1]. Although these conditions were historically associated with affluent, industrialized nations, the epidemiological landscape has shifted dramatically over recent decades. Low- and middle-income countries now bear over three-quarters of the global cardiovascular disease burden, and sub-Saharan Africa has experienced some of the most rapid increases in vascular disease mortality of any world region [2,3]. This transition is unfolding against a backdrop of stretched health systems, limited number of specialists and healthcare infrastructure, and high rates of undiagnosed hypertension, diabetes, and other vascular risk factors [4].

In Ghana, cardiovascular and cerebrovascular diseases have emerged as major contributors to hospital admissions and mortality over the past two decades. A recent systematic review and meta-analysis reported a pooled cardiovascular disease prevalence of 10.3% nationally, rising to 10.7% in hospital-based settings, with a steep rise in cardiac admissions documented over the same period [5,6]. Stroke is now the leading cause of neurological disability in the country, while hypertensive heart disease, heart failure, and ischemic heart disease account for a growing proportion of medical ward admissions at both teaching and district hospitals [7,8,9,10]. Despite this growing burden, most published data are derived from tertiary referral centers, leaving the experience of district hospitals, which serve the majority of the population, poorly classified. Additionally, the contribution of extraneous factors such as climate change to the development of these vascular diseases remains inadequately characterized in Ghana.

Studies from temperate climates have shown that weather and seasonal conditions influence the risk of vascular disease. Cold temperature exposure triggers vasoconstriction, elevates blood pressure, increases blood viscosity and platelet aggregability, and heightens sympathetic nervous system activity, each of which can precipitate acute cardiovascular and cerebrovascular events [11]. Multi-country ecological studies have documented a winter excess in cardiovascular and stroke mortality, with the greatest absolute burden observed in lower-income settings [12]. Associations between high humidity and cerebrovascular hospitalization have been described in some tropical and subtropical settings, and frequent extreme rainfall events have independently been linked to elevated cardiovascular mortality across a broad range of climates [13,14]. The applicability of these findings to tropical West Africa is, however, uncertain as few of the studies included countries in the sub region.

Ghana generally experiences two main meteorological patterns: a hot, dry season with minimal rainfall and a cooler rainy season. However, the Greater Accra Region in southern Ghana exhibits a bimodal rainfall regime, with a major rainy season extending from April to July and a minor rainy season from September to November. Based on long-term meteorological observations, the Ghana Meteorological Agency (GMet) has characterized these periods as distinct climatic regimes in the Greater Accra Region, where the present study was conducted [15].

The physiological pathways linking climate to vascular diseases, including thermoregulatory stress, altered physical activity, and exposure to indoor air pollution from biomass cooking fuels, may operate differently in tropical and temperate regions [16,17]. Population-level physiological adaptation to consistently high ambient temperatures in tropical settings is thought to lower the threshold for cold-induced vasoconstriction and sympathetic activation relative to temperate populations [17]. Consequently, even modest inter-seasonal decreases in temperature may have important cardiovascular implications in tropical settings [17]. However, this hypothesis remains inadequately tested because of the limited evidence from sub-Saharan Africa, where populations are exposed to high ambient temperatures throughout the year. Addressing this evidence gap is urgent, as climate change is projected to intensify seasonal temperature and rainfall variability across the region [18].

This study addresses how meteorological seasonality maps unto vascular disease admission patterns, using 45 months of available electronic health records from the Ga North District Hospital in Accra, Ghana. We characterized the climate profiles of each season at this location and described the temporal and seasonal trends in cardiovascular and cerebrovascular admissions at this hospital. The impact of meteorological variables on vascular hospitalization independent of patient-level factors was also assessed.

By focusing on a district-level facility serving a mixed urban–peri-urban catchment population, this study contributes empirical evidence from tropical West Africa and identifies climate change as a potential factor warranting consideration in vascular disease prevention and healthcare planning efforts.

## Materials and Methods

### Study design

We conducted a hospital-based ecological time-series analysis using electronic health records from Ga North District Hospital, Accra, Ghana, which is a government-managed district facility serving a mixed urban and peri-urban catchment population in the Greater Accra Region. The hospital provides general medical, surgical, and obstetric care while acting as a referral center for surrounding smaller health facilities. Thus, data from the hospital can provide useful insights into disease patterns, healthcare utilization, and public health challenges in Greater Accra. The study covered 45 calendar months from February 2022 to August 2025, capturing a total of 8,308 patient admissions. Data was assessed on September 8, 2025.

### Data sources

Clinical data were extracted from the electronic health records system of the hospital and included date of admission, age, sex, primary discharge diagnosis, in-hospital outcome (survived or died), public health insurance enrolment status, and cost of treatment in Ghanaian cedis (GHS). Monthly climate data were obtained from the GMet, the national authority responsible for the collection, analysis, and archiving of meteorological observations in Ghana. GMet provided station-level records for the Accra synoptic station covering the study period, including maximum and minimum monthly temperature, relative humidity, total monthly rainfall, and number of rainy-days per month. Mean temperature was derived as the arithmetic mean of maximum and minimum monthly values. These data were linked to patient records by admission month and year. For the purpose of this study, the meteorological season of Greater Accra was classified into three, based on their unique characteristics. The three meteorological seasons of the Greater Accra Region were defined as Dry and Warm (November to March), Wet and Cool (June to October), and Wet and Warm (April to May). This classification was based on the average temperatures, rainfall and humidity values received from the GMet for the period of the study. The seasons were assigned to each patient admission from the institutional dataset.

### Vascular burden classification

Vascular burden was classified from the primary discharge diagnosis registered in the electronic health record, without consideration of secondary diagnoses or comorbidities. Each admission was assigned to one of three mutually exclusive categories based on the World Health Organization International Classification of Diseases (ICD) codes: cardiovascular (cardiac and other vascular conditions recorded as the principal diagnosis), cerebrovascular (stroke and related cerebrovascular conditions recorded as the principal diagnosis), or non-vascular (all other principal diagnoses). This primary-diagnosis approach ensured that all category counts summed exactly to total admissions (N = 8,308) with no patient counted in more than one vascular group.

### Exclusion criteria

A record containing incomplete data was excluded from the analysis.

#### Statistical analysis

Descriptive statistics were reported as median with interquartile range (IQR) for continuous variables and count with percentage for categorical variables. Differences in patient characteristics across vascular burden categories were assessed using the Kruskal–Wallis rank-sum test for continuous variables and Pearson chi-squared tests with Monte Carlo simulation (B = 2000) for categorical variables. The climate profile of each meteorological season was summarized using the median and interquartile range of the climate variables. Annual and monthly admission proportions were calculated as condition-specific admissions divided by total admissions in the corresponding period and expressed as percentages. Seasonal distributions of admissions were examined using chi-squared goodness-of-fit tests to assess whether admissions were evenly distributed across the three meteorological seasons. In-hospital case-fatality rates were compared across seasons using chi-squared tests.

Logistic regression was used to examine associations between climate exposures and the probability of admission for cardiovascular or cerebrovascular disease relative to all other admissions. Climate variables were standardized to z-scores at the monthly level and merged with individual patient records such that each patient was assigned the climate conditions corresponding to their month of admission. Univariate models were fitted separately for each of the six climate variables.

Meteorological season was not included in the multivariable models because it represents a composite categorization of the climatic conditions under investigation and therefore captures the same underlying environmental variation as the climate variables themselves. Including both season and climate variables in the same model would introduce substantial redundancy and potentially obscure the independent associations of specific climatic exposures with vascular admissions. Consequently, multivariable models were constructed using non-redundant climate variables demonstrating the strongest univariate associations, with adjustment for age (continuous, per year) and sex (male versus female).

All analyses were conducted in R version 4.5.3, with statistical significance defined as p < 0.05.

#### Ethical consideration

This study used routinely collected administrative data from which patient identifiers had been removed before analysis. Permission to access and use the data was obtained from the authorities of Ga North District Hospital. In accordance with institutional policy for analyses of anonymized routine administrative data, formal ethics committee review was not required.

## Results

### Patient characteristics

The Ga North District Hospital recorded 8,308 admissions across 45 calendar months. Of these, 427 (5.1%) were classified as cardiovascular admissions, 672 (8.1%) as cerebrovascular admissions, and 7,209 (86.8%) as other admissions. Patient characteristics are presented in Table 1. Cardiovascular patients were the oldest group (median age 56 years, IQR 45 to 67), followed by cerebrovascular patients (median 52 years, IQR 38 to 64), while other admissions were substantially younger (median 37 years, IQR 25 to 52; p < 0.001). Cardiovascular admissions were predominantly female (61%), while cerebrovascular admissions showed a more balanced but male-leaning distribution (48% male; p = 0.017 across categories). Public health insurance enrolment was highest among cardiovascular patients (52%), compared with 40% of cerebrovascular and 41% of other admissions (p < 0.001). In-hospital mortality was markedly higher in both vascular groups: 9.6% (41 of 427) among cardiovascular patients and 11.0% (74 of 672) among cerebrovascular patients, compared with 3.2% (230 of 7,209) among other admissions (p < 0.001). Treatment costs were also significantly higher for vascular patients, with a median of 150 GHS (IQR 65.5 to 460.0) for cardiovascular, 125 GHS (IQR 60.0 to 235.0) for cerebrovascular, and 86 GHS (IQR 41.0 to 153.0) for other admissions (p < 0.001).

**Table 1.** Patient demographic and clinical characteristics by vascular burden category.

| **Characteristic** | **Cardiovascular**<br>**N = 427<sup>1</sup>** | **Cerebrovascular**<br>**N = 672<sup>1</sup>** | **Other Admissions**<br>**N = 7,209<sup>1</sup>** | **p-value<sup>2</sup>** |
| --- | --- | --- | --- | --- |
| **Age (years)** | 56 (45, 67) | 52 (38, 64) | 37 (25, 52) | <0.001 |
| **Sex** |  |  |  | 0.017 |
| Female | 260 (61%) | 352 (52%) | 4,084 (57%) |  |
| Male | 167 (39%) | 320 (48%) | 3,125 (43%) |  |
| **Public Health Insurance enrollment** |  |  |  | <0.001 |
| Yes | 221 (52%) | 270 (40%) | 2,966 (41%) |  |
| No | 206 (48%) | 402 (60%) | 4,243 (59%) |  |
| **In-hospital mortality** |  |  |  | <0.001 |
| Yes | 41 (9.6%) | 74 (11.0%) | 230 (3.2%) |  |
| No | 386 (90.4%) | 598 (89.0%) | 6,979 (96.8%) |  |
| **Cost of treatment (GHS)** | 150 (65.5, 460.0) | 125 (60.0, 235.0) | 86 (41.0, 153.0) | <0.001 |

<sup>1</sup>Median n (Q1-Q3). <sup>2</sup>Kruskal-Wallis rank sum test; Pearson's Chi-squared test with p-value.

### Climate characterization by meteorological season

The climate data confirmed three meteorological seasons climatically discrete during the study period (Table 2). The Dry and Warm season recorded the highest maximum temperatures (median 33.4 °C, IQR 33.2 to 33.9 °C), the highest mean temperature (29.9 °C, IQR 29.5 to 30.1 °C), and the lowest rainfall volume (10.0 mm/month, IQR 2.1 to 40.3) and rainy-day frequency (2.0 days/month, IQR 1.0 to 4.0). The Wet and Cool season presented a strikingly different profile with maximum temperatures being the lowest of the three seasons (30.0 °C, IQR 28.6 to 30.9 °C), mean temperature fell to 27.2 °C (IQR 26.2 to 27.6 °C), and a high relative humidity (81.8%, IQR 80.8 to 82.5). Rainfall was moderate in volume (67.5 mm/month, IQR 33.0 to 89.8) but characterized by frequent rain events (11.0 rainy-days/month, IQR 7 to 12). The Wet and Warm season combined the highest total rainfall of the three seasons (176.6 mm/month, IQR 101.3 to 232.2) with temperatures intermediate between the other two seasons (mean 29.3 degrees C, IQR 29.0 to 29.4). Rainy-day frequency (9 days/month, IQR 8.0 to 10.0) was somewhat lower than in the Wet and Cool season, despite substantially higher total rainfall, indicating that Wet and Warm precipitation falls more intensely over fewer events.

**Table 2.** Monthly climate variable summary by meteorological season during the study period.

| Climate variable | Dry and Warm | Wet and Cool | Wet and Warm |
| --- | --- | --- | --- |
| Max temperature (°C) | 33.4 (33.2–33.9) | 30.0 (28.6–30.9) | 32.8 (32.6–33.1) |
| Min temperature (°C) | 26.3 (25.9–26.5) | 24.2 (23.5–24.7) | 25.7 (25.5–25.8) |
| Mean temperature (°C) | 29.9 (29.5–30.1) | 27.2 (26.2–27.6) | 29.3 (29.0–29.4) |
| Relative humidity (%) | 75.1 (73.8–76.6) | 81.8 (80.8–82.5) | 77.5 (77.2–78.7) |
| Total rainfall (mm) | 10.0 (2.1–40.3) | 67.5 (33.0–89.8) | 176.6 (101.3–232.2) |
| Rainy-days per month (n) | 2.0 (1.0–4.0) | 11.0 (7–12.0) | 9 (8.0–10.0) |

*Values are median (Q1-Q3). Climate data sourced from the Ghana Meteorological Agency (GMet).*

### Temporal annual trends in vascular admissions

Cardiovascular admissions as a proportion of all admissions increased substantially over the study period: 2.4% (n = 46) in 2022, rising to 4.2% (n = 98) in 2023, peaking at 7.1% (n = 184) in 2024, before settling at 6.5% (n = 99) in 2025. This represents nearly a threefold increase over three full years. Cerebrovascular admissions followed a more gradual but consistent upward trajectory: 7.0% (n = 131) in 2022, 7.0% (n = 161) in 2023, 8.5% (n = 220) in 2024, and 10.5% (n = 160) in 2025 (Fig 1).

**Fig 1.**
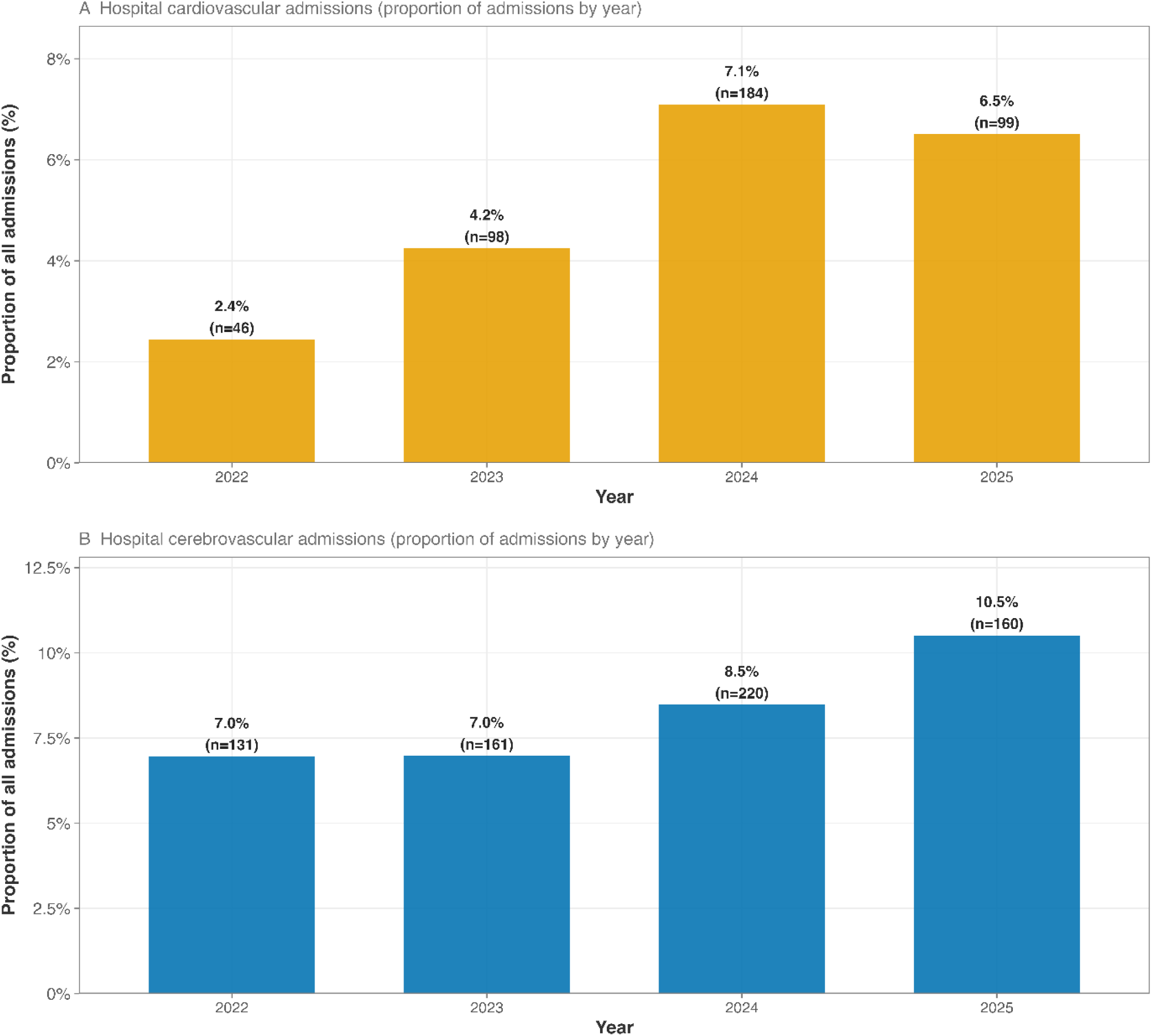
Yearly vascular disease admissions as a proportion of total hospital admissions.

### Temporal monthly trends in vascular admissions

Cardiovascular admissions showed moderate variation across the year, ranging from 4.4% to 6.9% of all admissions. The highest proportion occurred in July (6.9%), followed by August (5.2%), November (5.5%), and December (5.7%). The lowest proportion was recorded in June (4.4%), with similarly low values observed in January (4.5%) and April (4.6%). Overall, cardiovascular admissions tended to increase during the Wet and Cool season (June–October), particularly in July, before remaining relatively elevated towards the end of the year (Fig 2).

**Fig 2.**
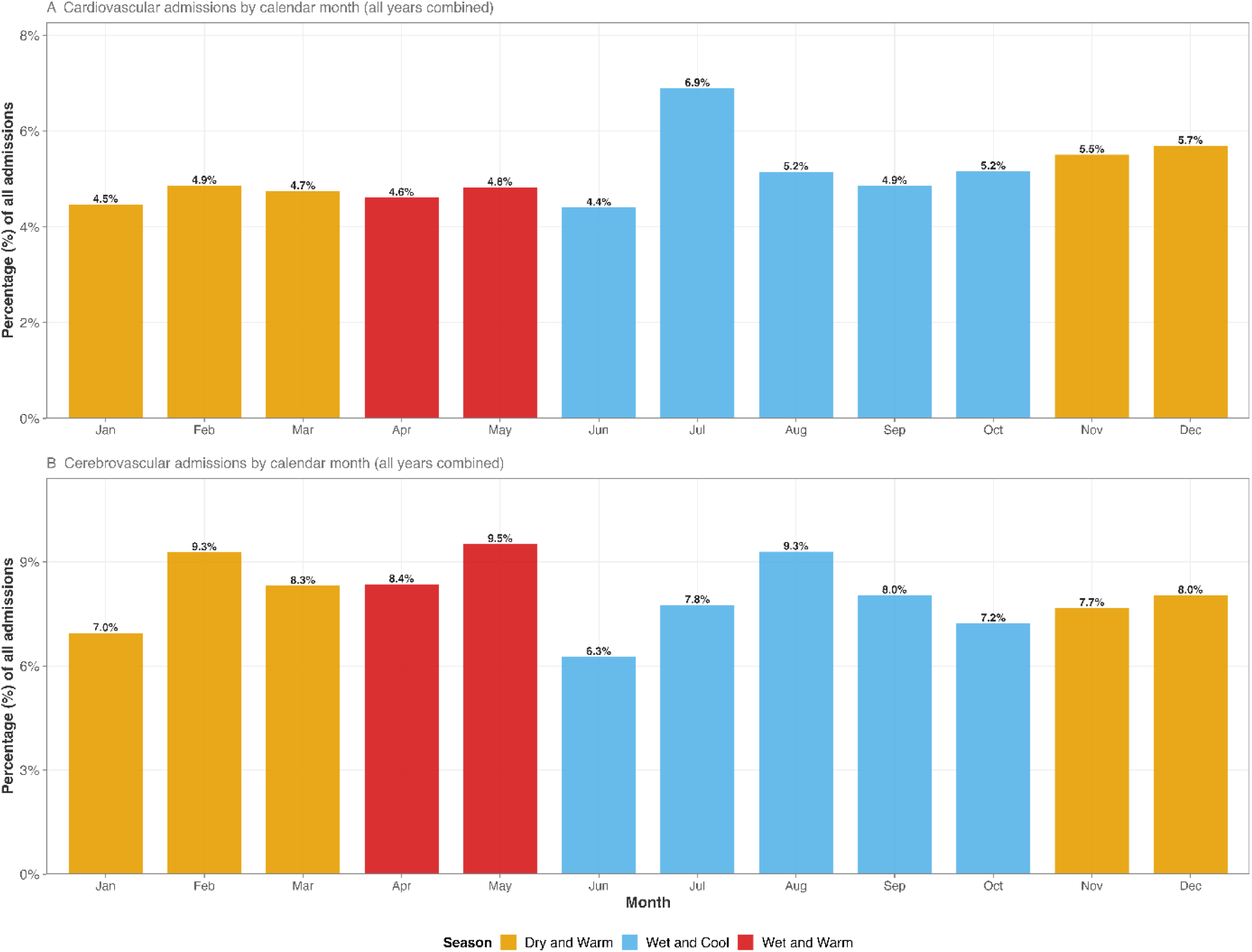
Monthly vascular disease admissions as a proportion of total hospital admissions, pooled across all study years.

Cerebrovascular admissions on the other hand demonstrated greater month-to-month variation than cardiovascular admissions, ranging from 6.3% to 9.5% of all admissions. The highest proportions were observed in May (9.5%), February (9.3%), and August (9.3%), indicating peaks during both warm and cool periods of the year. The lowest proportion occurred in June (6.3%), followed by January (7.0%) and October (7.2%). Unlike cardiovascular admissions, cerebrovascular admissions did not exhibit a single dominant peak but rather multiple peaks distributed across different seasons.

### Seasonal distribution of vascular admissions

Both cardiovascular and cerebrovascular admissions were disproportionately concentrated in the Wet and Cool season (Fig 3). Among cardiovascular patients, 46.1% (n = 197) presented during the Wet and Cool season, 38.2% (n = 163) during the Dry and Warm season, and 15.7% (n = 67) during the Wet and Warm season. The pattern was similar for cerebrovascular patients: 42.1% (n = 283) in the Wet and Cool season, 39.0% (n = 262) in the Dry and Warm season, and 18.9% (n = 127) in the Wet and Warm season. Chi-squared goodness-of-fit tests confirmed that these distributions were highly non-uniform for both conditions (cardiovascular: chi-squared = 63.87, p < 0.001; cerebrovascular: chi-squared = 63.99, p < 0.001) thus, rejecting the null hypothesis of equal distribution across seasons.

**Fig 3.**
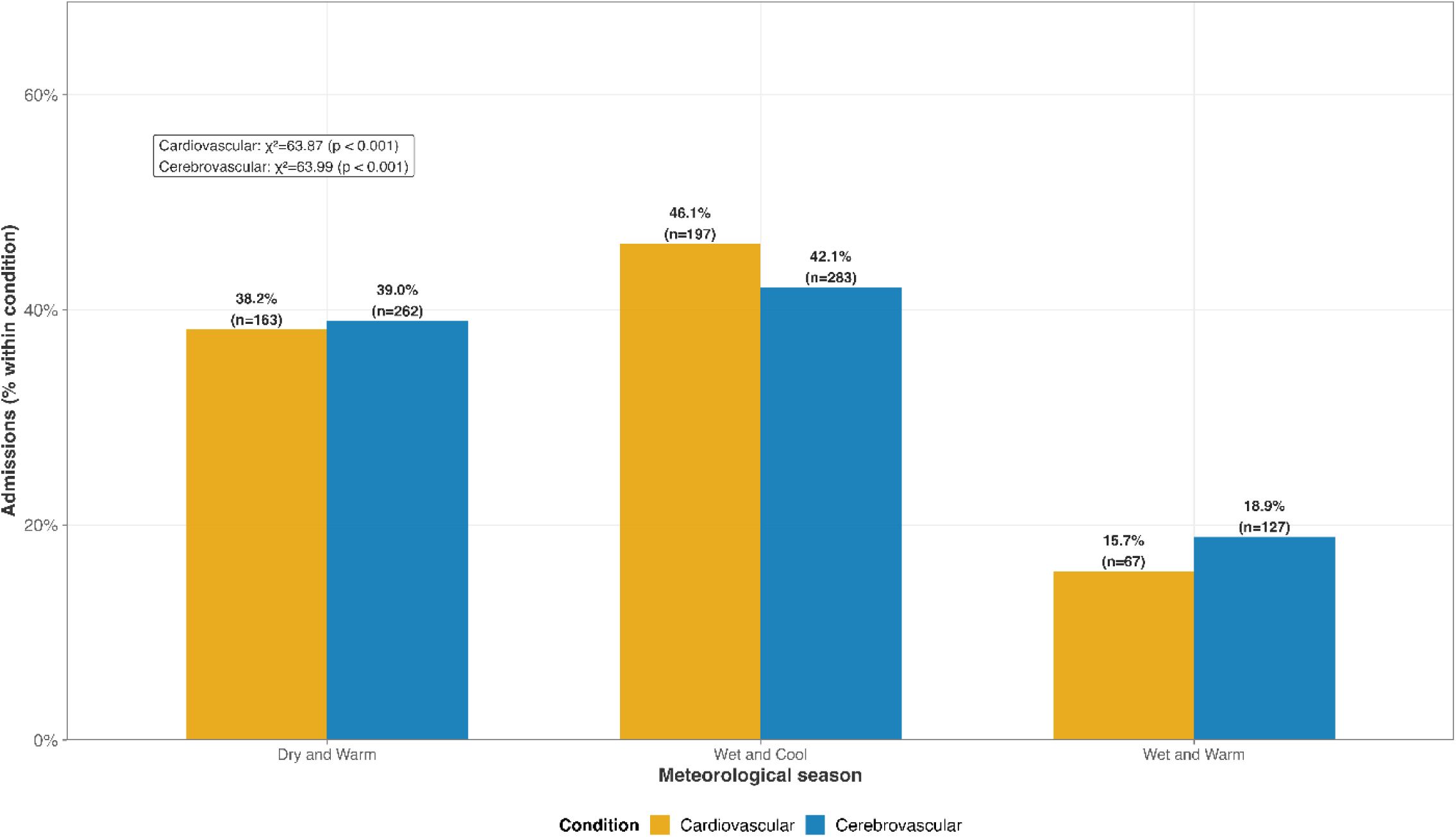
Distribution of vascular disease admissions by meteorological season.

### In-hospital mortality by season

In-hospital case-fatality rates by meteorological season are shown in Fig 4. For cardiovascular patients, case-fatality was 10.4% (17 of 163) in the Dry and Warm season, 9.1% (18 of 197) in the Wet and Cool season, and 9.6% (6 of 67) in the Wet and Warm season (chi-squared = 0.21, p = 0.890). For cerebrovascular patients, case-fatality was highest in the Dry and Warm season at 13.0% (34 of 262), compared with 10.2% (29 of 283) in the Wet and Cool season and 8.7% (11 of 127) in the Wet and Warm season (chi-squared = 1.92, p = 0.357). Neither comparison reached statistical significance.

**Fig 4.**
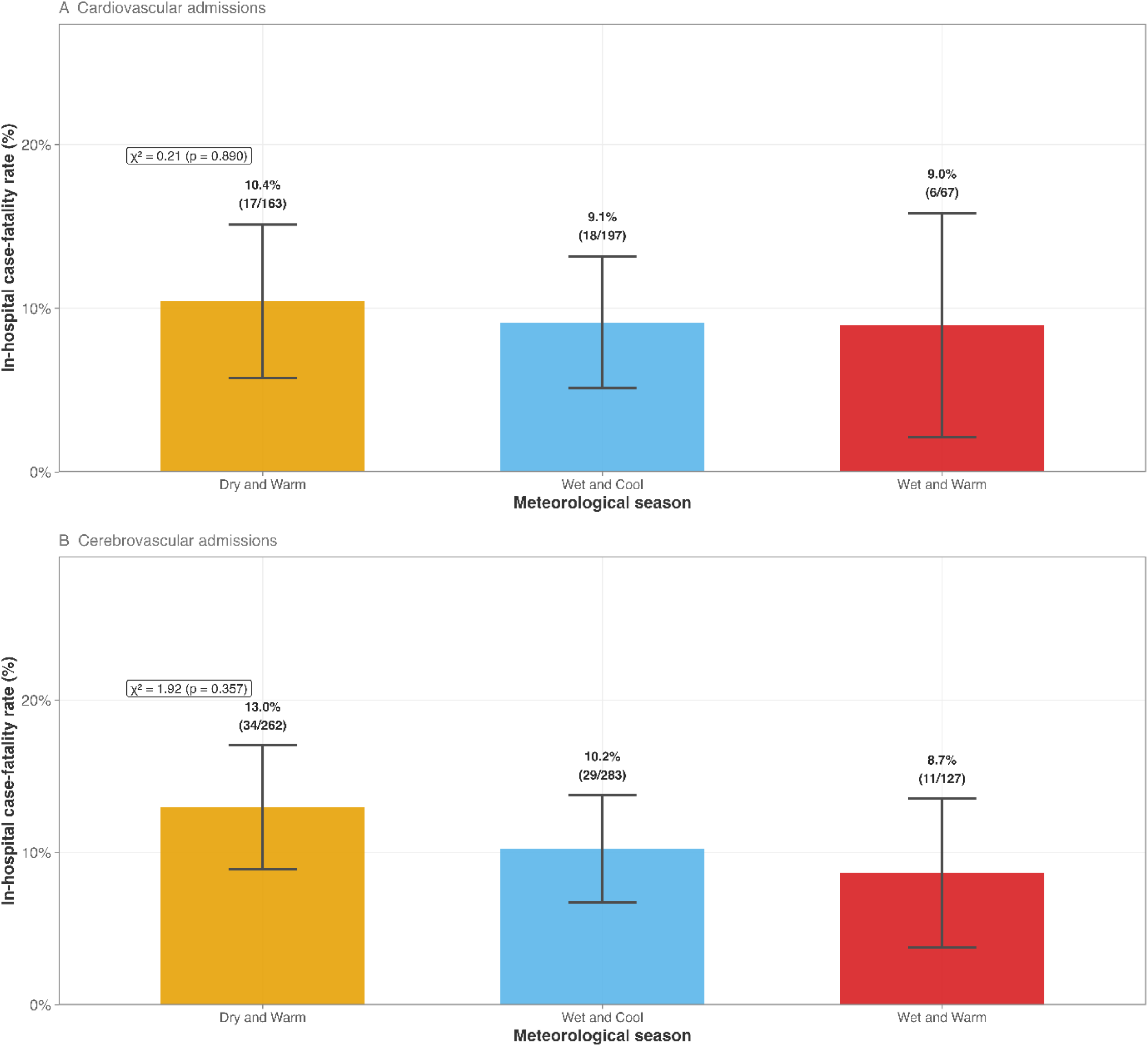
In hospital case fatality by meteorological season.

### Predictors of cardiovascular admission

For cardiovascular admissions (Table 3), only rainy-day count was significantly associated with the outcome in univariate analysis (OR 1.12 per 1-SD increase, 95% CI 1.01 to 1.24, p = 0.028); temperature, humidity, and total rainfall were not significant individually. In the multivariable model adjusting for all non-redundant climate variables simultaneously alongside age and sex, two rainfall-related variables emerged as independent predictors with divergent directions. Higher total rainfall volume was associated with lower odds of cardiovascular admission (OR 0.72, 95% CI 0.61 to 0.85, p < 0.001), while greater rainy-day frequency was associated with higher odds (OR 1.73, 95% CI 1.38 to 2.18, p < 0.001). Mean temperature and relative humidity were not independently significant after mutual adjustment. Each additional year of age increased the odds of cardiovascular admission by 4% (OR 1.04, 95% CI 1.04 to 1.05, p < 0.001), and sex was not a significant predictor.

**Table 3.** Logistic regression: Predictors of cardiovascular admission.

| Predictor | OR (95% CI) | p-value |
| --- | --- | --- |
| **Univariate analyses** |  |  |
| Max temperature, per 1 SD | 0.92 (0.84–1.01) | 0.090 |
| Min temperature, per 1 SD | 1.02 (0.92–1.13) | 0.726 |
| Mean temperature, per 1 SD | 0.95 (0.87–1.05) | 0.334 |
| Relative humidity, per 1 SD | 1.05 (0.95–1.16) | 0.349 |
| Total rainfall, per 1 SD | 0.92 (0.83–1.02) | 0.116 |
| Rainy-days per month, per 1 SD | 1.12 (1.01–1.24) | 0.028 |
| **Multivariable model** |  |  |
| Mean temperature (°C) (per 1 SD) | 1.05 (0.85–1.30) | 0.662 |
| Relative humidity (%) (per 1 SD) | 0.80 (0.61–1.04) | 0.091 |
| Total rainfall (mm) (per 1 SD) | 0.72 (0.61–0.85) | < 0.001 |
| Rainy-days per month (n) (per 1 SD) | 1.73 (1.38–2.18) | < 0.001 |
| Age, per 1 year | 1.04 (1.04–1.05) | < 0.001 |
| Sex: male vs female | 0.94 (0.77–1.16) | 0.572 |

OR = odds ratio; CI = confidence interval; SD = standard deviation. Climate predictors standardised (z-scored) at monthly level and merged to patient-level records. The multivariable model includes all non-redundant climate variables simultaneously (mean temperature, relative humidity, total rainfall, and rainy-days per month), adjusted for age and sex.

### Predictors of cerebrovascular admission

For cerebrovascular admissions (Table 4), none of the six climate variables were significantly associated with the outcome in univariate analysis. In the multivariable model including all non-redundant climate variables simultaneously with age and sex, no climate variable reached statistical significance: mean temperature OR 1.09 (95% CI 0.92 to 1.29, p = 0.342), relative humidity OR 1.01 (95% CI 0.82 to 1.24, p = 0.944), total rainfall OR 0.95 (95% CI 0.84 to 1.07, p = 0.393), and rainy-days OR 1.10 (95% CI 0.92 to 1.32, p = 0.293). Older age remained a strong independent predictor (OR 1.03 per year, 95% CI 1.03 to 1.03, p < 0.001), and male sex was independently associated with higher odds of cerebrovascular admission (adjusted OR 1.30, 95% CI 1.10 to 1.53, p = 0.002).

**Table 4.** Logistic regression: Predictors of cerebrovascular admission.

| Predictor | OR (95% CI) | p-value |
| --- | --- | --- |
| **Univariate analyses** |  |  |
| Max temperature, per 1 SD | 1.01 (0.93–1.09) | 0.799 |
| Min temperature, per 1 SD | 1.04 (0.96–1.13) | 0.297 |
| Mean temperature, per 1 SD | 1.02 (0.94–1.11) | 0.579 |
| Relative humidity, per 1 SD | 1.00 (0.92–1.08) | 0.955 |
| Total rainfall, per 1 SD | 0.99 (0.91–1.07) | 0.748 |
| Rainy-days per month, per 1 SD | 1.01 (0.94–1.10) | 0.724 |
| **Multivariable model** |  |  |
| Mean temperature (°C) (per 1 SD) | 1.09 (0.92–1.29) | 0.342 |
| Relative humidity (%) (per 1 SD) | 1.01 (0.82–1.24) | 0.944 |
| Total rainfall (mm) (per 1 SD) | 0.95 (0.84–1.07) | 0.393 |
| Rainy-days per month (n) (per 1 SD) | 1.10 (0.92–1.32) | 0.293 |
| Age, per 1 year | 1.03 (1.03–1.03) | < 0.001 |
| Sex: male vs female | 1.30 (1.10–1.53) | 0.002 |

*OR = odds ratio; CI = confidence interval; SD = standard deviation. Climate predictors standardised (z-scored) at monthly level and merged to patient-level records. The multivariable model includes all non-redundant climate variables simultaneously (mean temperature, relative humidity, total rainfall, and rainy-days per month), adjusted for age and sex.*

## Discussion

This study provides a four-year characterization of cardiovascular and cerebrovascular hospitalizations at a district facility in Ghana, documenting their rising burden, their strong and statistically significant seasonal concentration, and the divergent roles of meteorological and patient-level factors in predicting each condition. The findings make four principal contributions. First, the GMet climate data confirmed that three meteorological seasons at this location are substantially distinct environments, providing a defensible empirical basis for seasonal analysis in a setting where such data are rarely linked to health records. Second, both cardiovascular and cerebrovascular admissions increased markedly over the study period, with cardiovascular cases nearly tripling in three years, signalling a rapidly escalating burden at the district level. Third, both conditions were concentrated in the Wet and Cool season with remarkable statistical consistency, as evidenced by chi-squared values exceeding 63 and p-values below 0.001 for both, a finding that has direct implications for health system planning. Fourth, the factors underlying seasonal concentration appeared to differ between the two conditions. Cardiovascular risk was partly associated with a measurable climate factor (rainy-day count), whereas cerebrovascular risk was associated primarily with individual demographic characteristics.

The rising trajectory of vascular admissions at the study facility is consistent with the broader epidemiological transition documented across urban sub-Saharan Africa and aligns with regional projections of growing cardiovascular and cerebrovascular burden as populations age and urbanize [3,19]. Whereas there was a steady rise in cerebrovascular cases over the study period, the cardiovascular increase was particularly steep, more than doubling between 2022 and 2023 and peaking at nearly three times the 2022 proportion by 2024. Whether this reflects a genuine worsening of population-level cardiovascular health, improved case ascertainment, increased health-seeking behavior at the district level, or some combination of these cannot be determined from administrative data alone. Nevertheless, recent GMet reports indicate that Ghana experienced persistently high temperatures during the 2023–2025 period (Supplementary Fig). Temperatures were particularly elevated in 2024 and 2025, with 2025 ranking among the warmest years observed over the past 35 years, further supporting evidence of a sustained warming trend in the country [20]. With continuously elevated temperatures, the proportion of clinical, pharmacological, and bed capacity required for vascular disease management at this facility could grow substantially, a trajectory that is likely to accelerate given projected demographic and climatic trends.

In relation to total monthly hospital admissions, cardiovascular cases peaked in July, whereas cerebrovascular cases peaked in February, May and August. February and March are the hottest months, May marks a warm transitional period during the major rainy season, and August is typically the coolest month with a period of reduced rainfall within the major rainy season [21,22]. Thus, increases in cerebrovascular admissions were observed during periods corresponding to both warm and cool temperature extremes in the annual temperature cycle in Accra. For seasonal concentration of both vascular cases, there was unequal distribution of admissions across the meteorological seasons. The variation observed for both cardiovascular and cerebrovascular diseases were statistically significant (p < 0.001), providing strong evidence against the null hypothesis of a uniform meteorological season distribution. The Wet and Cool season accounted for approximately 44% of all hospital admissions. Within this period, 46.1% of cardiovascular disease admissions and 42.1% of cerebrovascular disease admissions were recorded, indicating a significant seasonal concentration of vascular disease admissions beyond that expected under a uniform seasonal distribution. Conversely, the Wet and Warm season, despite generating the highest rainfall volume of the year, accounts for a disproportionately small share of vascular admissions relative to its duration. This pattern, replicated for both conditions and across all four study years, provides observational evidence that the climatic profile of the Wet and Cool season, rather than rainfall volume per se, is an important contributor to vascular admission concentration at this facility. The GMet data clarify what makes the Wet and Cool season climatically distinct. It is the only season in the annual cycle of Accra that combines the lowest temperatures, the highest relative humidity, and frequent rainfall. The mean temperature of 27.2°C represents a drop of approximately 2.7°C relative to the Dry and Warm season and 2.1°C relative to the Wet and Warm season. These inter-seasonal differences may appear modest by temperate standards, but they are physiologically meaningful in a population whose thermoregulatory systems are calibrated to consistently high ambient temperatures. Population-level adaptation to high ambient temperatures is thought to lower the threshold for cold-induced vasoconstriction and sympathetic nervous system activation, such that temperature drops that would be physiologically trivial in a temperate population can trigger significant hemodynamic responses in a tropical one [17]. High humidity during the Wet and Cool season compounds this effect by impairing evaporative heat loss and increasing the thermoregulatory burden on the cardiovascular system, independent of the temperature contribution [13,23]

The simultaneous inclusion of all non-redundant climate variables in the multivariable model revealed a more comprehensive assessment of climate-related cardiovascular risk than analyses based on individual predictors alone. Total rainfall volume and rainy-day frequency emerged as independent predictors with strikingly divergent effects. Each standard deviation increase in total monthly rainfall was associated with 28% lower odds of cardiovascular admission (OR 0.72, p < 0.001), while each standard deviation increase in rainy-day frequency was associated with 73% higher odds (OR 1.73, p < 0.001). The persistence of both associations after adjustment for other climatic factors suggests that rainfall amount and rainfall frequency may reflect different patterns of meteorological exposure that are independently associated with cardiovascular admissions. These observations align with findings reported by He et al., whose two-stage time-series study involving 645 locations worldwide showed that frequent rainfall events were linked to increased cardiovascular mortality independent of total precipitation volume [14]. The similar pattern observed between the two studies provides further support for the hypothesis that the cardiovascular effects of rainfall are more closely related to rainfall frequency than the cumulative rainfall amount over a given period. Periods in which rainfall is concentrated into a few intense events may cause less sustained disruption to daily behavioural activities than months characterised by recurrent rainfall. Frequent rainy-days may contribute to cardiovascular risk through several pathways, including reduced opportunities for outdoor physical activity, decreased sun exposure, and increased exposure to indoor air pollution from prolonged biomass fuel combustion in inadequately ventilated homes [16,14]. Heavy but infrequent rainfall does not sustain these exposures across the month in the same way. Mean temperature and relative humidity did not independently predict cardiovascular admission after adjustment for rainfall variables. These findings suggest that rainfall frequency and rainfall patterns may be more strongly associated with cardiovascular hospitalisation than temperature or humidity within this study population.

The stark divergence between the climate sensitivity of cardiovascular and cerebrovascular admissions is one of the most informative findings of this study. Despite sharing a common seasonal concentration pattern and comparable in-hospital mortality, the two conditions demonstrated differing patterns of association after statistical adjustment. Cardiovascular admissions were climate sensitive. Rainy-day count remained a significant predictor after adjustment for age, sex, and other non-redundant climatic variables. Cerebrovascular admissions, on the other hand, were not significantly predicted by the climate variables when age and sex were included in the model. The presence of seasonal concentration in the absence of significant climatic predictors suggests that factors co-varying with season but not captured by the meteorological variables included in this study, may contribute to cerebrovascular admission patterns. Interestingly, male sex emerged as an independent predictor of cerebrovascular admission in the adjusted model, a finding consistent with the global epidemiology of stroke incidence, which shows higher rates in men across most age groups, attributed to higher prevalences of hypertension, smoking, alcohol consumption, and diabetes [24,25]. The Stroke Investigative Research and Educational Network (SIREN) case-control study, conducted in Ghana and Nigeria, identified hypertension as the dominant modifiable stroke risk factor in these populations and documented that men carry a higher burden of overall modifiable risks [10]. The convergence of our findings with the SIREN data suggests that individual-level characteristics may play a more prominent role in cerebrovascular admissions than the climatic variables examined in this study, while cardiovascular admissions were additionally shaped by the environmental conditions of their month of presentation.

The predominantly female cardiovascular population at this facility (61% female) is consistent with the high burden of hypertensive heart disease and heart failure among women in urban sub-Saharan Africa [26]. Hypertension is more prevalent among urban women than men in many parts of the region, and heart failure secondary to hypertension is a leading cardiac diagnosis. The lower odds of cardiovascular admission for male patients in the adjusted model (OR 0.92, though non-significant) suggests that the sex distribution here reflects genuine epidemiological patterns rather than health-seeking differences alone.

In-hospital mortality though slightly higher in both vascular groups (9.6% cardiovascular and 11.0% cerebrovascular) in the Dry and Warm months, did not differ significantly. The absence of a significant seasonal pattern may reflect limited statistical power resulting from the small number of deaths within each season. Overall, the mortality rates were broadly consistent with reported case-fatality rates for vascular conditions in district and regional hospitals across Ghana and Nigeria [27]. These mortality levels may also reflect the broader healthcare challenges previously described in the region, including limited access to early specialist care, diagnostic imaging, and intensive care [27].

These findings collectively have practical implications for climate-resilient health system planning in tropical low- and middle-income countries. District hospitals in West Africa manage the majority of acute vascular presentations in their catchment areas, and our findings suggest that these facilities should systematically anticipate heightened cardiovascular and cerebrovascular demand during the major rainy season. Operational adaptations could include seasonal upscaling of cardiovascular medication stocks, deployment of additional nursing staff during the Wet and Cool period, and community-level health messaging on physical activity maintenance and indoor air quality management on frequent rainy-days. As climate change is expected to intensify rainfall variability and inter-seasonal temperature contrasts across sub-Saharan Africa, [18] the seasonal patterns observed in this study may also change in timing or magnitude, highlighting the importance of continued meteorological surveillance in climate-adaptive health planning.

### Limitations

This study was unable to adjust for individual-level confounders, including hypertension, diabetes, smoking status, physical activity, and body mass index, all of which may influence vascular admission risk and exhibit seasonal variation. Consequently, residual confounding cannot be excluded. In addition, the study was conducted at a single district-level facility, which may limit the generalizability of the findings to other settings. Due to administrative challenges beyond the control of the hospital and authors, hospital records for January 2022 and September–December 2025 were unavailable for analysis, resulting in incomplete coverage of the study period. Furthermore, lagged effects of climatic exposures were not examined, and therefore the study may not have captured delayed influences of meteorological conditions on vascular admissions occurring in subsequent weeks or months.

## Conclusion

The Wet and Cool season, characterized by lower temperatures, higher humidity, and frequent rainfall, was associated with a higher concentration of both cardiovascular and cerebrovascular hospital admissions at the Ga North District Hospital. After adjustment for age and sex, rainy-day frequency remained an independent predictor of cardiovascular admissions, suggesting that environmental and behavioural factors associated with recurrent rainfall may contribute to cardiovascular risk. In contrast, although cerebrovascular admissions exhibited a similar seasonal concentration pattern, they were not independently associated with the climatic variables examined and were more strongly associated with patient-level characteristics, particularly older age and male sex. In-hospital mortality was highest during the Dry and Warm season for both conditions but the differences were not statistically significant.

These findings provide empirical evidence from sub-Saharan Africa on the relationship between meteorological conditions and vascular disease burden. The study also demonstrates the feasibility of integrating electronic health records with national meteorological data to establish a scalable framework for climate-sensitive health surveillance. In the context of ongoing epidemiological transition and accelerating climate change, such evidence may help inform climate-responsive public health strategies and strengthen the resilience of health systems in low- and middle-income countries.

## Supporting Information

**S1 Fig.**
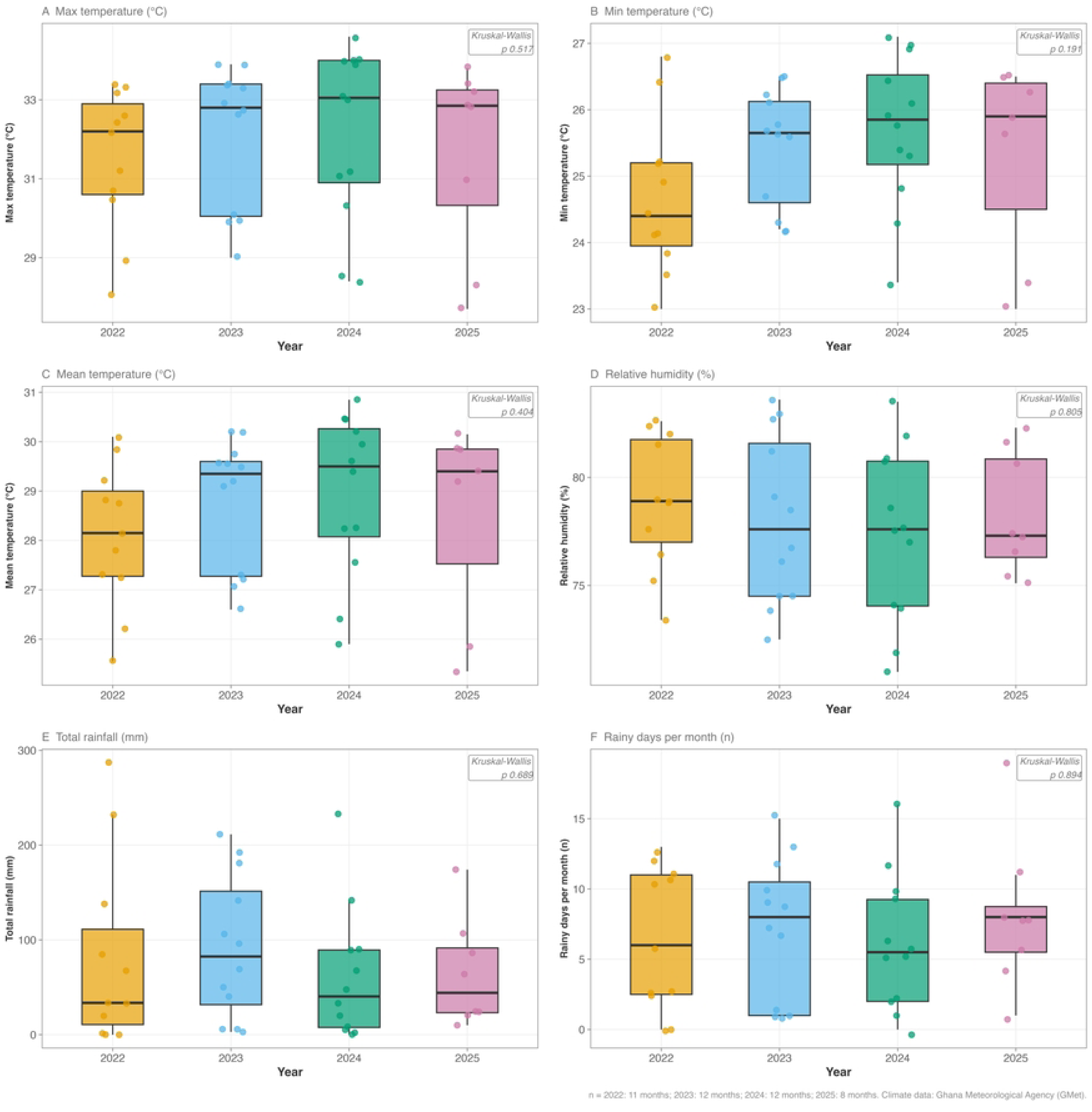
Annual Distribution of Climate Variables.

## Data Availability

The dataset analyzed in this study is held by Ga North District Hospital and the Ghana Health Service. Data sharing is subject to institutional data governance approval. Requests should be directed to the corresponding author.

